# Smoking behaviour in Swiss schoolchildren: the LuftiBus in the school survey

**DOI:** 10.1101/2020.05.29.20116525

**Authors:** Rebeca Mozun, Cristina Ardura-Garcia, Carmen C. M. de Jong, Myrofora Goutaki, Jakob Usemann, Florian Singer, Philipp Latzin, Claudia E. Kuehni, Alexander Moeller

**Author notes:** These authors contributed equally to the work (Claudia E. Kuehni, Alexander Moeller). **Correspondence to:** Alexander Moeller Universitäts-Kinderspital Zürich – Eleonorenstiftung Steinwiesstrasse 75, CH-8032 Zurich, Switzerland.

## Abstract

**Background:** Smoking is a major preventable cause of morbidity and mortality. We assessed smoking behaviour in 6-17-year-olds. In 13-17-year-olds, we studied risk factors for smoking and compared the frequency of respiratory problems between those who smoked and those who did not.

**Methods:** We used data from *LuftiBus in the school*, a school-based survey of the respiratory health of children carried out 2013-2016 in the canton of Zurich, Switzerland. Participants were asked about cigarette, shisha, and electronic smoking device (ESD) smoking, and respiratory symptoms and diseases. We studied the association between smoking and risk factors using logistic regression.

**Results:** We included 3488 schoolchildren with data on active smoking. Five percent of the 6-12-year-olds (90 of 1905) had smoked occasionally (<once/week). Among 13-17-year-olds (N=1583), 563 (36%) had smoked occasionally of whom 414 smoked ESDs, 409 shishas, and 276 cigarettes. Among 54 (3%) 13-17-year-olds who smoked frequently (≥once/week), 41 (76%) smoked cigarettes, and 22% of 15-17-year-olds (104/477) had smoked all three products. Smoking was more common in adolescents who were male, lived in rural areas, and whose mother (adjusted OR 1.7, 95% CI 1.3 - 2.3) or father (aOR 1.5, 95% CI 1.2 – 1.9) smoked. Respiratory symptoms in the past year such as rhinitis, dyspnoea, and wheeze were more common in adolescents who smoked frequently or occasionally than in never smokers.

**Conclusion:** Smoking of shishas and ESDs is popular among Swiss adolescents and often combined with smoking cigarettes. Structural smoking prevention strategies should include all forms of smoking including shishas and ESDs.

## Introduction

Smoking and nicotine consumption are major preventable causes of morbidity and mortality worldwide^1^. In Switzerland, 9500 people die directly or indirectly due to smoking each year^2,3^. According to surveillance reports from the Swiss Federal Office of Health, over one-quarter of the Swiss population aged 15 years and older smoke regularly^3,4^. Prevalence of tobacco smoking has declined among Swiss adolescents in the last years^5^, but smoking behaviours are evolving. Shishas, also known as water pipes, and especially electronic smoking devices (ESDs) such as e-cigarettes, have become increasingly popular among adolescents. ESDs are currently the most common form of smoking in US adolescents^6^. ESD smoking anytime within the last 30-days increased from 12% in 2017 to 21% in 2018 among high school students in the US^6^”^8^. These new types of smoking are not harmless^9,1^°. Adolescent and adult ESD users have increased risk of upper and lower respiratory symptoms and animal studies show damage to bronchial epithelia. Longitudinal studies suggest as well that adolescents who begin smoking ESDs are likely to switch to smoking cigarettes in the future, the so-called gateway effect^n^. It is thus important to prevent children and adolescents from starting to smoke any type of product^12,13^.

Data on smoking behaviour of Swiss adolescents are scarce. Some countries including Switzerland do not yet regulate the advertising, sale, and flavouring of ESDs, which is likely to encourage children and adolescents to start smoking^13^. Better understanding the characteristics of adolescents who smoke and the type of products they prefer could help the design of better, more effective preventive strategies. We assessed active smoking of cigarettes, shishas, and ESDs in school-aged children from the canton of Zurich in Switzerland and, among adolescents, we identified risk factors for smoking and compared the frequency of respiratory symptoms and diseases between those who had and had not smoked.

## Methods

### Study design and setting

*LuftiBus in the school* is a cross-sectional school-based study conducted between 2013 and 2016 in the canton of Zurich, Switzerland. All schools in the canton were invited to participate (N = 490). The school directors decided whether to take part in the study, and with which classes. A bus visited 37 schools that took part in the study. The ethics committee of the canton of Zurich approved the study (KEK-ZH-Nr: 2014–0491). The study was funded by *Lunge Zürich*, Switzerland.

### Study procedures (recruitment and selection)

Before the school visit, the children’s parents were asked to complete a questionnaire and gave informed consent. During the visit of the bus to the school, the children were interviewed using a short questionnaire. The fieldworkers interviewing the children were unaware of the answers on the parental questionnaire, and parents remained unaware of children’s answers. Parental and children’s questionnaires contained validated questions about respiratory symptoms and diseases from the International Study of Asthma and Allergies in Childhood and the Leicester Respiratory Cohorts^14,15^. The fieldworkers measured children’s standing height and body weight without shoes in the bus.

### Study population and inclusion criteria

We analysed data about active smoking of cigarettes, shishas, or ESDs from participating children aged 6 to 17 years. We assessed frequency of active smoking in the whole sample of 6–17-year-olds. We studied factors associated with active smoking among those aged 13 to 17 years, whom we refer to as adolescents, among whom occasional or frequent smoking was more common.

### Frequency of smoking cigarettes, shishas, and ESDs

Participants were asked questions about active smoking of three different products: Have you ever smoked cigarettes (commercial or self-rolled), Have you ever smoked shisha, and Have you ever smoked e-cigarettes or e-shisha (with or without nicotine)? For simplicity we refer to e-cigarettes or e-shishas as ESDs. We categorized answers relating to the smoking frequency of cigarettes, shishas, and ESDs into three ordered groups: never smoked, smoked occasionally (“once or twice” or “less than once per week”), and smoked frequently (“at least once per week” or “everyday”). We defined “any smoking” as smoking any of the three products occasionally or frequently. The original formulation of the questions can be found in the online material (E-Table 1 and E-Table 2).

### Sociodemographic exposures

We collected information about sex, country of birth of the participant, sports, physical activity, maternal smoking during pregnancy, and parental country of origin, history of asthma, and education from the parental questionnaire (online E-Table 1 and E-Table 2). Information on current parental smoking was obtained from the children’s questionnaire because it had fewer missing values and correlated well with parental smoking as reported in the parental questionnaire. BMI z-scores were calculated using reference tables for the Swiss population^16^. We defined obesity as a BMI z-score ≥ 2. We used the Swiss socioeconomic position index (Swiss-SEP) from the Swiss national cohort as an area-based measure of socioeconomic status^17,18^. This measure is based on data about rent per square meter, education and occupation of households’ heads, and household crowding, and ranges from 0 (lowest) to 100 (highest)^18^. We matched the geocodes of the addresses of our participants to the closest geocode of the Swiss socioeconomic position index dataset for the canton of Zurich. If the address or address geocode of a participant was missing, we assigned the mean socioeconomic position index of the participant’s school. The degree of urbanization of the municipality of the schools was categorized as large urban, small urban, or rural area according to the Swiss federal office of statistics classification^19^. A municipality was categorised as large urban area if at least half of the population lived in high-density clusters, as rural area if more than half of the population lived in rural grid cells, and as small urban area if less than half of the population lived in rural grid cells and less than half lived in a high-density cluster.

### Respiratory symptoms and diseases

Information on respiratory symptoms in the past 12 months and respiratory diseases was taken from the children’s questionnaire. We assessed respiratory symptoms in the past 12 months that included cough apart from colds, coughing more than peers, rhinitis apart from colds, a dry mouth when waking-up in the morning as a proxy for nocturnal mouth breathing, dyspnoea, wheeze, and exercise induced wheeze. We also assessed two respiratory diseases, hay fever and having received a diagnosis of asthma from a doctor ever in their life (online E-Table 1 and E-Table 2).

### Statistical analysis

We described smoking frequency in the whole study sample of 6–17-year-olds.

Frequent smoking started at age 13 or older, so we analysed risk factors and respiratory problems in those aged 13–17 years. We compared proportions of risk factors between categories of frequency of smoking using p-values for a trend for binary and categorical variables and ANOVA for continuous variables. To assess independent effects of risk factors for smoking, we used logistic regression. We selected age, sex, socioeconomic position index, urbanisation, and paternal and maternal smoking a priori as explanatory variables based on the literature^4,20,21^. in a sensitivity analysis, we also included paternal and maternal education in the model.

We studied crude proportions of respiratory symptoms and diseases as reported by the adolescents. In a sensitivity analysis, we also studied the association between respiratory symptoms and frequency of active smoking of any product using logistic regression and adjusting for potential confounders: age, sex, and hay fever or asthma diagnosis. We assumed that reporting of respiratory problems by adolescents is less prone to information bias than reporting by parents since adolescents are probably more aware of their own symptoms, especially during exercise or night time, than their parents. We excluded observations with missing data. Information on missing values can be found in the online material (online E-Table 5). We used the software STATA (Version 14, StataCorp., College Station, TX, USA) for statistical analysis.

We used the Strengthening the Reporting of Observational Studies in Epidemiology (STROBE) guidelines for reporting in cross-sectional studies^22^.

## Results

3488 participants had consent and information on active smoking of cigarettes, shishas, or ESD. We focused our attention mainly in the subsample of 1583 adolescents (13–17 years), of whom 100 (6%) were obese (Table 1). Most, 912 (58%), lived in small urban areas of the canton of Zurich, and 1140 (87%) were born in Switzerland, although half of their parents originally came from countries other than Switzerland. Mean socioeconomic position index was 64.7 (SD 10.0). The fathers of 467 adolescents (30%) and the mothers of 313 (20%) were current smokers. Maternal smoking during pregnancy was reported by 117 (9%).

**TABLE 1:** Characteristics of participants aged 13 to 17 years by smoking status (N= 1583).

|  | Total |  | Frequency of smoking for any product |  |  |  |  |  | p trend |
| --- | --- | --- | --- | --- | --- | --- | --- | --- | --- |
|  |  |  | Never |  | Occasional |  | Frequent |  |  |
|  | n | (%) | n | (%) | n | (%) | n | (%) |  |
| Sex, male | 793 | (50) | 426 | (44) | 331 | (59) | 36 | (66) | <0.001 |
| Age in years; mean (SD) | 14.5 | (0.9) | 14.3 | (0.9) | 14.7 | (0.9) | 15.4 | (0.9) | <0.001<sup>a</sup> |
| Obesity | 100 | (6) | 48 | (5) | 47 | (8) | 5 | (9) | 0.008 |
| Sport apart from school | 919 | (71) | 598 | (71) | 287 | (68) | 34 | (77) | 0.606 |
| Physical activity |  |  |  |  |  |  |  |  |  |
| Very active | 277 | (21) | 153 | (18) | 107 | (25) | 17 | (40) | 0.001 |
| Moderately active | 807 | (62) | 539 | (65) | 246 | (59) | 22 | (51) |  |
| Not very active | 211 | (16) | 140 | (17) | 67 | (16) | 4 | (9) |  |
| Child country of birth, CH | 1140 | (87) | 730 | (87) | 367 | (86) | 43 | (98) | 0.504 |
| Father country of origin, CH | 709 | (55) | 469 | (57) | 216 | (51) | 24 | (55) | 0.123 |
| Mother country of origin, CH | 669 | (52) | 439 | (53) | 209 | (50) | 21 | (48) | 0.251 |
| Urbanisation degree |  |  |  |  |  |  |  |  |  |
| Large urban area | 564 | (36) | 315 | (33) | 223 | (40) | 26 | (48) | 0.005 |
| Small urban area | 912 | (58) | 590 | (61) | 296 | (53) | 27 | (48) |  |
| Rural area | 107 | (7) | 61 | (6) | 44 | (8) | 2 | (4) |  |
| Socioeconomic position index; mean (SD) | 64.7 | (10.0) | 64.9 | (10.2) | 64.1 | (9.8) | 65.8 | (9.5) | 0.177<sup>a</sup> |
| Highest paternal education |  |  |  |  |  |  |  |  |  |
| Elementary school | 102 | (12) | 55 | (11) | 41 | (13) | 6 | (17) | 0.020 |
| Professional school | 309 | (37) | 176 | (36) | 116 | (38) | 17 | (49) |  |
| Business/technical school | 138 | (17) | 84 | (17) | 50 | (16) | 4 | (11) |  |
| Teacher education / University | 288 | (34) | 182 | (36) | 98 | (32) | 8 | (23) |  |
| Highest maternal education |  |  |  |  |  |  |  |  |  |
| Elementary school | 132 | (15) | 74 | (15) | 52 | (17) | 6 | (15) | 0.033 |
| Professional school | 393 | (46) | 223 | (44) | 143 | (46) | 27 | (68) |  |
| Business/technical school | 115 | (13) | 69 | (14) | 44 | (14) | 2 | (5) |  |
| Teacher education / University | 216 | (25) | 137 | (27) | 74 | (24) | 5 | (13) |  |
| Paternal current smoking |  |  |  |  |  |  |  |  |  |
| No | 1085 | (70) | 699 | (74) | 357 | (64) | 29 | (57) | <0.001 |
| Yes, sometimes | 197 | (13) | 112 | (12) | 78 | (14) | 7 | (14) |  |
| Yes, often | 270 | (17) | 134 | (14) | 121 | (22) | 15 | (30) |  |
| Maternal current smoking |  |  |  |  |  |  |  |  |  |
| No | 1235 | (80) | 794 | (84) | 411 | (75) | 30 | (58) | <0.001 |
| Yes, sometimes | 140 | (9) | 82 | (9) | 52 | (9) | 6 | (12) |  |
| Yes, often | 173 | (11) | 69 | (7) | 88 | (16) | 16 | (31) |  |
| Maternal smoking during pregnancy | 117 | (9) | 65 | (8) | 47 | (11) | 6 | (14) | 0.027 |
| Parental history of asthma |  |  |  |  |  |  |  |  |  |
| None | 1122 | (87) | 725 | (87) | 359 | (86) | 38 | (86) | 0.621 |
| Maternal | 105 | (8) | 66 | (8) | 37 | (9) | 2 | (5) |  |
| Paternal | 62 | (5) | 40 | (5) | 18 | (4) | 4 | (9) |  |
| Both parents | 7 | (1) | 3 | (0) | 4 | (1) | 0 | (0) |  |

Any product: cigarettes, shishas or electronic smoking devices. Occasional smoking: once-twice or <once/week.
Frequent smoking: ≥once/week. Obesity: BMI z-score ≥2. The urbanization degree of the location of the schools was categorized according to the Swiss federal office of statistics classification. SD: standard deviation. CH: Switzerland. P trend: test for trend across ordered groups. <sup>a</sup>ANOVA: analysis of variance and covariance

### Use of cigarettes, shishas, and ESDs in the whole study sample

Smoking of each product increased with age (Figure 1). In the younger subsample of 612-year-olds (N = 1905), 90 (5%) had smoked any product occasionally. Below the age of 10 years, 14 out of 919 children had smoked (2%). Ten out of 289 children aged 10 years (3%) had smoked, among whom 5 children had smoked cigarettes, 2 ESDs, and 3 shishas. Among 11-year-olds, 17 out of 299 had smoked (6%), 6 of whom had smoked cigarettes, 6 ESDs, and 6 shishas. Among those aged 12 years, 50 out of 398 children (13%) had smoked, 17 of whom had smoked cigarettes, 24 ESDs, and 22 shishas.

**FIGURE 1:**
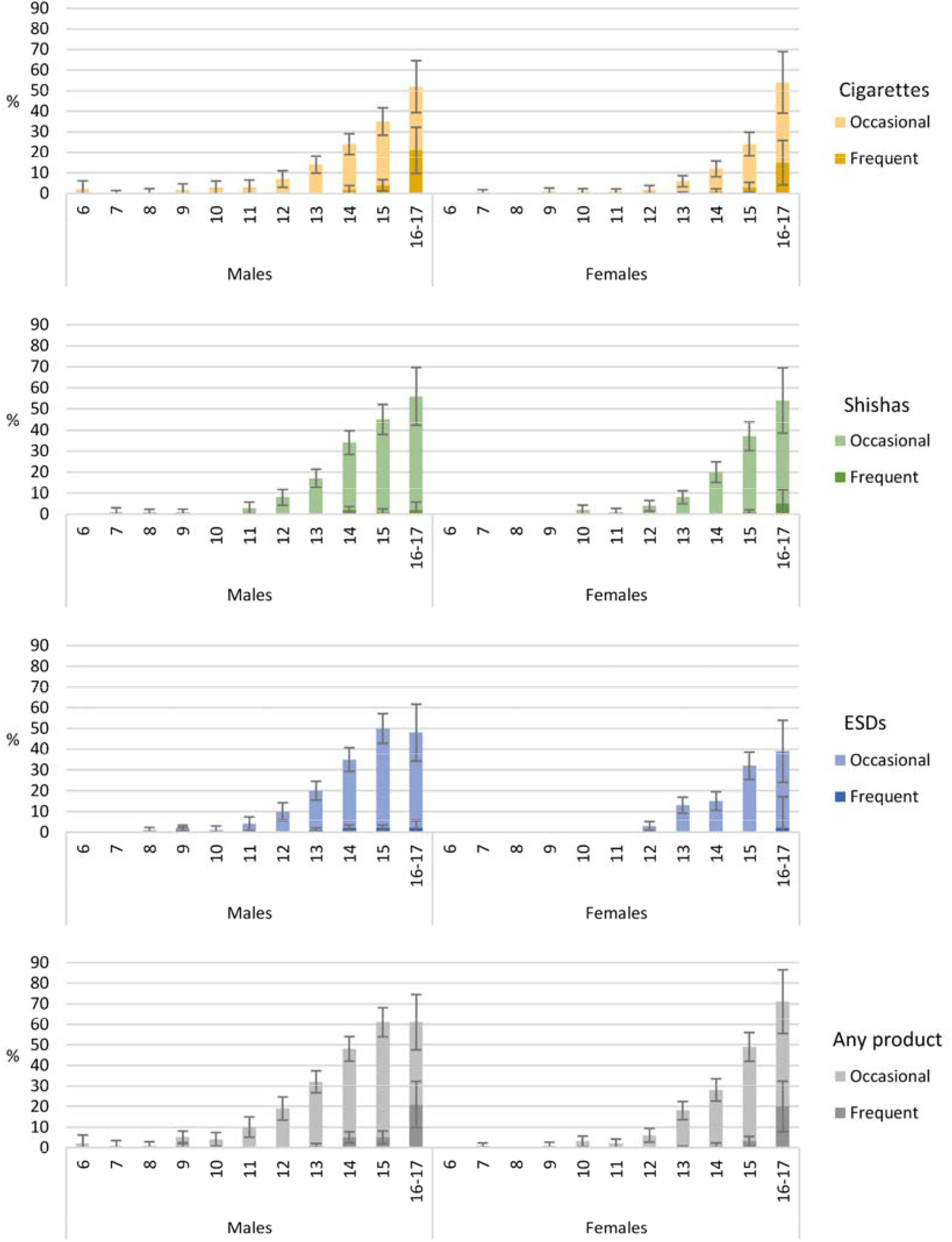
Prevalence of active smoking among adolescents aged 6 to 17 years, stratified by age and sex (N = 3488). Any product: cigarettes, shishas or ESDs. ESDs: electronic smoking devices. Occasional smoking: once-twice or<once/week. Frequent smoking: ≥ once/week. Whiskers represent 95% confidence intervals.

Among adolescents, 966 (61%) had not smoked, 563 (36%) had smoked any product occasionally, and 54 (3%) any product frequently. From age 13 to 15 years, more boys than girls smoked occasionally (42% vs 28%) or frequently (3% vs 1%) (p< 0.01 for the sex difference). However, among 16–17-year-olds, 51% of girls smoked occasionally and 20% frequently, whereas 40% of the boys smoked occasionally and 21% frequently (p = 0.55 for the sex difference).

Regarding the type of product smoked, out of the 563 adolescents who smoked occasionally, 414 adolescents (74%) smoked ESDs, 409 (73%) shishas, and 276 (49%) cigarettes. Among the 54 adolescents who smoked frequently, 41 (76%) smoked cigarettes, 11 (20%) ESDs, and 10 (19%) shishas. Adolescents often combined smoking cigarettes, shishas, and ESDs (Figure 2). Among 15–17-year-olds, 121 (25%) had smoked ESDs and cigarettes, 147 (31%) shishas and ESDs, and 104 (22%) had smoked all three.

**FIGURE 2:**
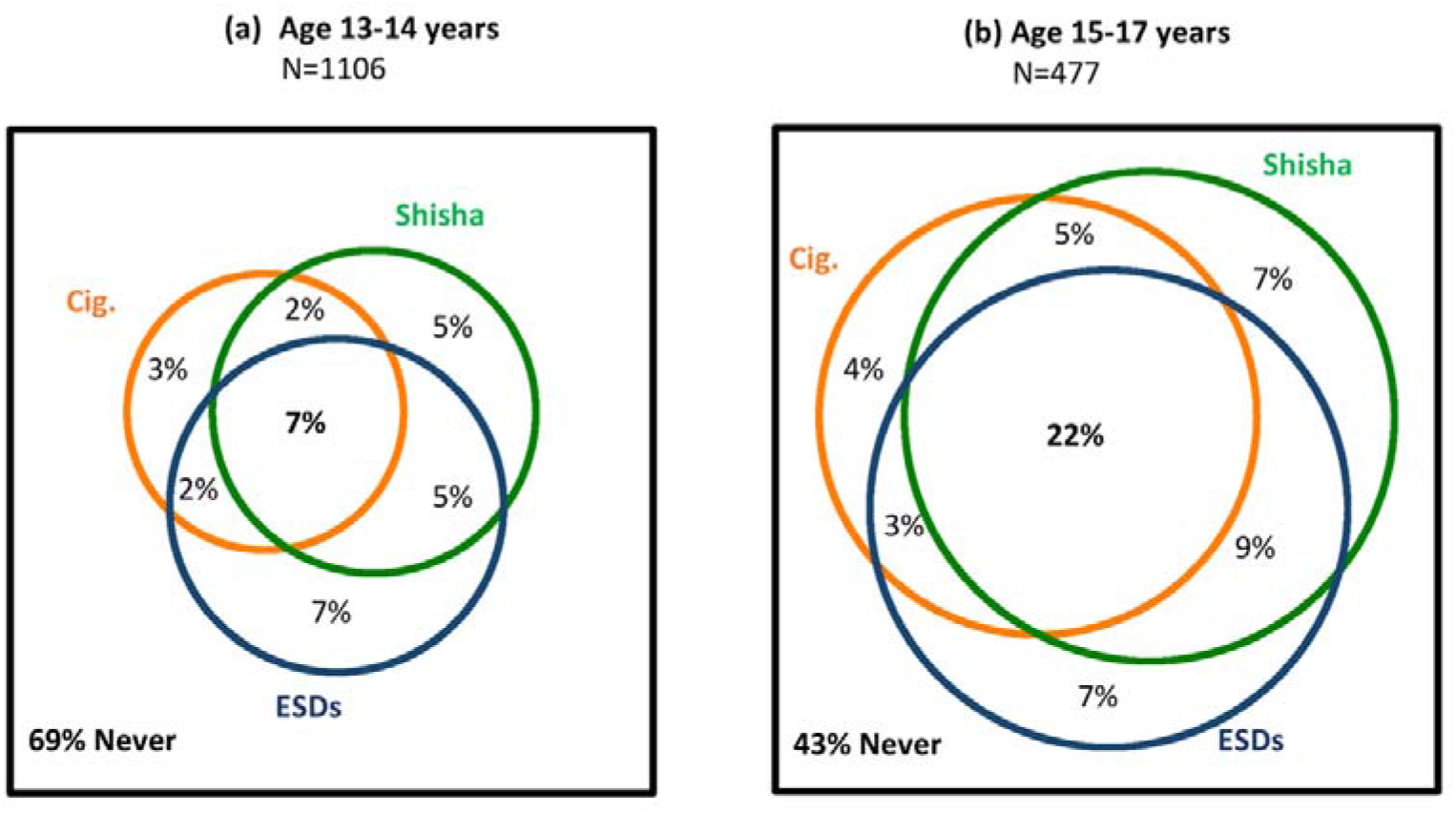
Venn diagrams showing the overlap between occasional or frequent active smoking of cigarettes, shishas and electronic smoking devices in adolescents aged (a) 13–14 and (b) 15–17 years. ESDs: electronic smoking devices.

### Sociodemographic risk factors for adolescent active smoking of any product

Participants who reported any smoking more often were male (adjusted odds ratio 2.1 95% confidence interval 1.7 – 2.6), older (aOR 2.0 per year increase of age, 95% CI 1.8 – 2.3), and had a mother (aOR 1.7, 95% CI 1.3 – 2.3) or father (aOR 1.5, 95% CI 1.2 – 1.9) who smoked (Figure 3, online E-Table 4). Any smoking was more common in rural (aOR 1.8, 95% CI 1.2 – 2.9) and large urban areas (aOR 1.2, 95% CI 0.9–1.6) than in small urban areas. We found no association with socioeconomic position index (aOR 1.0 per decile increase, 95% CI 0.9 –1.1), or with paternal and maternal education (online ETable 5).

**FIGURE 3:**
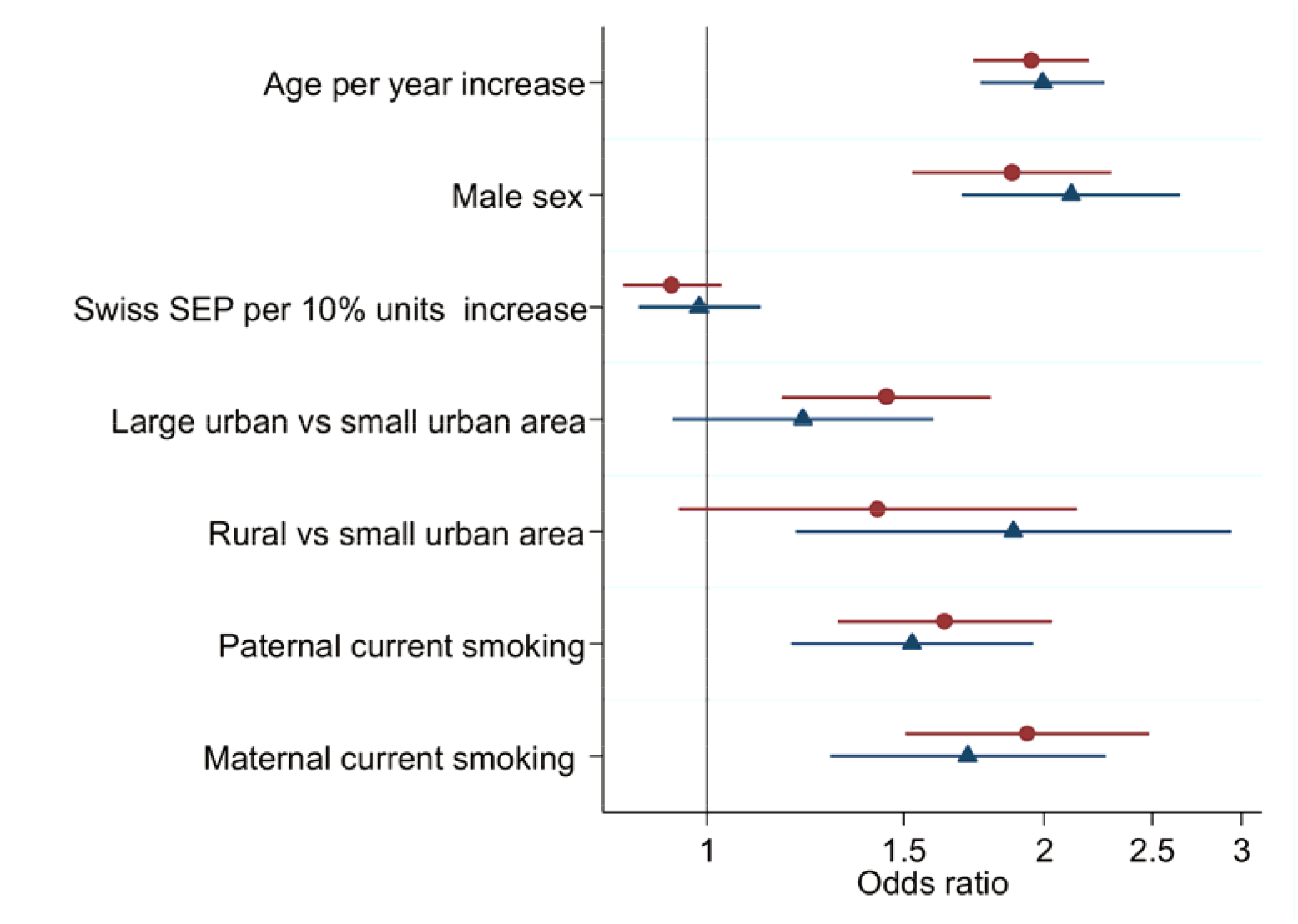
Unadjusted (circle) and adjusted (triangle) odds ratios for the association between sociodemographic factors and adolescent reported occasional or frequent smoking (N = 1527). The adjusted model includes all the variables in the figure. A table with the adjusted and unadjusted odds ratios and confidence intervals can be found in the supplementary material (E-Table 4).

### Respiratory symptoms in adolescents by frequency of active smoking of any product

Smokers had more respiratory symptoms in the past 12 months than nonsmokers, even if the smoking was only occasional (Table 2). This was also true when adjusting for potential confounders, with similar point estimates when adjusting for age, sex, and asthma or hay fever, and also when adjusting for paternal and maternal smoking (online E-Table 6). Upper airway respiratory symptoms were more common among adolescents who smoked. Rhinitis apart from colds was more often reported by frequent smokers (44%) and occasional smokers (32%) than by never smokers (29%). Similarly, 17% of adolescents who smoked frequently had a dry mouth when waking up, as did 13% of those who smoked occasionally and 9% of nonsmokers. Rhinitis apart from colds and having a dry mouth when waking up were more common in occasional smokers (aOR for rhinitis 1.1, 95% CI 0.9–1.4; aOR for dry mouth 1.6, 95% CI 1.1–2.3,) and frequent smokers (aOR for rhinitis 1.9, 95% CI 1.1–3.5; aOR for dry mouth 2.6, 95% CI 1.2–5.8) than in never smokers adjusting for age, sex, and hay fever.

**TABLE 2:** Self-reported respiratory symptoms and diseases in adolescents aged 13–17 years by frequency of self-reported smoking of any type.

|  | Total<br>N=1583 |  | Frequency of smoking any product |  |  | p |
| --- | --- | --- | --- | --- | --- | --- |
|  |  |  | Never<br>N=966 | Occasional<br>N=563 | Frequent<br>N=54 |  |
|  | n | (%) | n (%) | n (%) | n (%) |  |
| **Respiratory symptoms in the past year** |  |  |  |  |  |  |
| Cough apart from colds | 430 | (27) | 250 (26) | 162 (29) | 18 (34) | 0.112 |
| Cough more than others | 64 | (4) | 36 (4) | 24 (4) | 4 (8) | 0.261 |
| Rhinitis apart from colds | 472 | (30) | 273 (29) | 175 (32) | 24 (44) | 0.024 |
| Dry mouth often when waking-up | 162 | (10) | 85 (9) | 68 (13) | 9 (17) | 0.011 |
| Dyspnoea | 317 | (20) | 181 (19) | 120 (22) | 16 (30) | 0.046 |
| Wheeze | 156 | (10) | 77 (8) | 73 (13) | 6 (12) | 0.003 |
| Exercise triggered wheeze | 225 | (14) | 119 (12) | 94 (17) | 12 (22) | 0.004 |
| **Respiratory diseases** |  |  |  |  |  |  |
| Hay fever | 386 | (24) | 228 (24) | 143 (25) | 15 (28) | 0.348 |
| Asthma diagnosis ever | 163 | (11) | 97 (10) | 59 (11) | 7 (13) | 0.609 |

Any product: cigarettes, shishas or electronic smoking devices. Occasional smoking: once-twice or <once/week.
Frequent smoking: ≥once/week. P is the p value for a test for trend for each respiratory symptom or disease across the ordered categories of frequency of smoking.

Lower airway respiratory symptoms were also more frequent among smokers. Wheeze was more common in frequent (12%) or occasional smokers (13%) than in never smokers (8%). Adolescents reported exercise-triggered wheeze more frequently than wheeze, and exercise-triggered wheeze was also more common in frequent (22%) or occasional smokers (17%), than in never smokers (12%). Similarly, dyspnoea was more often reported by frequent (30%) and occasional (22%) smokers than never smokers (19%). When adjusting for age, sex, and asthma diagnosis, when compared to never smokers occasional smokers had more wheeze (aOR 2.1, 95% CI 1.5–3.1), exercise induced wheeze (aOR 1.9, 95% CI 1.3–2.6) and dyspnoea (aOR 1.4, 95% CI: 1.0–1.8). The adjusted odds of wheeze were twice as high in frequent smokers compared to never smokers (aOR 2.0, 95% CI 0.8 – 5.4), exercise induced wheeze (aOR 3.2, 95% CI 1.5–6.7), and dyspnoea (aOR 2.2, 95% CI 1.1–4.5). Compared to never smokers, adolescents tended to have more cough apart from colds if they smoked occasionally (aOR 1.3, 95% CI 1.0–1.7) or frequently (aOR 1.7, 95% CI 0.9–3.2), and to cough more if they smoked occasionally (aOR 1.4, 95% CI 0.8–2.4) or frequently (aOR 3.0, 95% CI 0.9–9.5), adjusting for confounders.

## Discussion

A significant proportion of adolescents smoked and often combined smoking cigarettes, shishas, and ESDs. Occasional smoking of shishas and ESDs was more popular than smoking cigarettes. Most of those who smoked frequently were 16–17 years old and smoked cigarettes, which could reflect higher nicotine dependency.

More adolescents who smoked had a mother or father who smoked. Adolescents who smoked frequently or occasionally had more respiratory symptoms in the past 12 months including rhinitis apart from colds, a dry mouth when waking up in the morning, dyspnoea, and wheeze and exercise induced wheeze than never smokers.

### Strengths and limitations

The school-based settings in which we collected information using structured questionnaires with validated questions on respiratory problems are ideal for studying smoking behaviour in school-aged children. School directors decided whether to participate in the study and with which classes, independently of the smoking status of the students, which may have reduced selection bias. Still, our sample was not randomly selected and may not be representative of the whole population of school-aged children living in the canton of Zurich. We did not objectively assess nicotine consumption by cotinine measurements, and response bias could have led to an underestimation of smoking among adolescents. However, we believe that response bias may have been reduced by children’s answers not being available to their parents. We did not collect information on brand names or composition of ESDs smoked, and future studies should collect this information to allow more specific consumption monitoring and health risk assessments^9^.

### Comparison with previous studies

Smoking was common among adolescents in our study. Prevalence of frequent smoking of cigarettes in Switzerland was lower than in Germany, Italy, and France, but higher than in Denmark, England, and Sweden according to reports of the Health Behaviour in School aged Children (HBSC) surveys^23^. Similarly, prevalence of cigarette smoking in Swiss adults seems below the European average^24,25^. However, Jakob et al. showed that the cigarette consumption estimated by aggregate sales data is larger than survey-based estimates for Switzerland^24^. In that study, the discrepancy between actual and reported cigarette consumption was higher in Switzerland (46%) than France (7%). Underreporting could be a reason why smoking prevalence seems to be relatively low in Switzerland despite its loose tobacco regulation. The US national tobacco surveys showed that the prevalence of ESD use in the past 30 days increased while cigarette smoking decreased between 2011–2018 among high school students aged 13–17 years^6^. The marked increase in use of ESDs among high school students in the US coincided with a steep increase in the sales of certain ESD brands that are now also available in the Swiss market^6^”^8^. Studies of ESD and shisha use in European adolescents report different prevalence measures, include different age groups, and were done in different years making comparison between countries challenging.

Paternal and maternal smoking were independently associated with active smoking among adolescents in our study. This is in line with previous studies^20,26^. A study from Germany found that parental cigarette smoking was a risk factor for ESD smoking in adolescents^21^. Peer behaviour also plays an important role in adolescent smoking behaviour^20,21^.Adolescents living in rural areas smoked more often than those in urban areas independently of their family’s socioeconomic position and parental smoking status. A recent study from the US reported a smaller reduction in the prevalence of adolescent cigarette smoking in rural than in urban areas, implying that preventive strategies may be less strict in those areas^28^. Social drivers of ESD and shisha smoking and their relationship with cigarette smoking deserve further investigation^9^.

Smoking any product was associated with respiratory symptoms in our study, even if the smoking was occasional. A study in adolescents from Chile also showed that frequent active smoking of cigarettes is associated with respiratory symptoms^29^. A large population-based cohort study from the US found significant decline in lung function even among light adult cigarette smokers^30^. Population-based surveys in Asia and North America also found increased risks of upper and lower respiratory symptoms among adolescents who smoke ESDs^10,31^”^33^. McConnell et al. found higher risk of chronic cough and bronchitis among adolescents who had smoked ESDs even after adjusting for potential confounders^31^.

### Implications for public health and for research

Use of ESDs by adolescents is not safe and can have adverse consequences for personal and public health^34^. With or without nicotine, ESDs expose the respiratory airways to toxic and irritating agents such as propylene glycol, glycerol, and flavourings. ESDs smoking affects nasal and bronchial epithelia by impairing ciliary function and altering gene expression^10^. Furthermore, a recent meta-analysis showed that adolescents and young adults who had smoked ESDs ever in their life were subsequently more likely to start smoking cigarettes than those who had never smoked ESDs (pooled aOR 3.5, 95% CI 2.4 – 5.2)^11^

We need to protect adolescents from every type of smoking, and this may require adapting preventive policies. Switzerland signed the WHO framework convention on tobacco control in 2004, but has still not ratified it^35^. Swiss preventive strategies that involve regulation of tobacco affordability, advertising, and smoking in public spaces are less strict than those of other European countries^24^. Exposure to tobacco advertising, in particular, has been shown to increase the likelihood of smoking initiation among adolescents^8^,^36^,^37^. Currently, marketing of ESD brands often targets adolescents by advertising through social media and offering a variety of candy flavours^28^. Regulation of ESDs was absent in Switzerland and sales were free of age limitation during our study^9^, though this is likely to change in coming years due to new federal legislation currently under study. The need as well to regulate quality control of the ingredients and flavouring of ESDs has been highlighted by research^13^. The composition of ESDs varies greatly, and lung injury associated with ESDs containing tetrahydrocannabinol caused several deaths in the US^38^. Yet in spite of harms already documented, the long-term effects of ESD smoking on lung function and the full range of associated respiratory symptoms are unclear and require further research^10^.

## Conclusion

Occasional smoking of shishas and ESDs is popular among Swiss adolescents, who often combine smoking shishas, ESDs, and cigarettes. We encourage structural preventive strategies that focus not only on traditional cigarettes, but also on shishas and ESDs.

## Data Availability

The LuftiBus in the school dataset is available on reasonable request by contacting Alexander Moeller by

## Acknowledgements

We thank schools, children and their families for taking part in the study. We thank fieldworkers of the LuftiBus in the school study for their technical support during the study. We thank Christopher Ritter (Institute of Social and Preventive Medicine, University of Bern) for his editorial contributions.

## Author contributions

Alexander Moeller, Claudia E. Kuehni and Philipp Latzin conceptualised and designed the study. Alexander Moeller supervised data collection. Rebeca Mozun analysed the data and drafted the manuscript. Cristina Ardura-Garcia, Carmen C. M. de Jong, Myrofora Goutaki, Jakob Usemann and Florian Singer supported the statistical analysis and gave input for interpretation of the data. All authors critically revised and approved the manuscript.

